# Validation and clinical discovery demonstration of a real-world data extraction platform

**DOI:** 10.1101/2023.02.21.23286092

**Authors:** Amanda Nottke, Sophia Alan, Elise Brimble, Anthony B. Cardillo, Lura Henderson, Hana E. Littleford, Susan Rojahn, Heather Sage, Jessica Taylor, Lisandra West-Odell, Alexandra Berk

## Abstract

**Objective:** To validate and demonstrate the clinical discovery utility of a novel patient-mediated, medical record collection and data extraction platform developed to improve access and utilization of real-world clinical data.

**Methods:** Clinical variables were extracted from the medical records of consented patients with metastatic breast cancer. To validate the extracted data, case report forms completed using the structured data output of the platform were compared to manual chart review for 50 patients. To demonstrate the platform’s clinical discovery utility, we assessed associations between time to distant metastasis (TDM) and tumor histology, molecular type, and germline *BRCA* status in the platform-extracted data of 194 patients.

**Results:** The platform-extracted data had 97.6% precision (91.98%–100% by variable type) and 81.48% recall (58.15%–95.00% by variable type) compared to manual chart review. In our discovery cohort, the shortest TDM was significantly associated with metaplastic (739.0 days) and inflammatory histologies (1,005.8 days), HR-/HER2- molecular types (1,187.4 days), and positive *BRCA* status (1,042.5 days) as compared to other histologies, molecular types, and negative *BRCA* status, respectively. Multivariable analyses did not produce statistically significant results, but the average TDMs are reported.

**Discussion:** The platform-extracted clinical data are precise and comprehensive. The data can generate clinically-relevant insights.

**Conclusion:** The structured real-world data produced by a patient-mediated, medical record-extraction platform are reliable and can power clinical discovery.

## INTRODUCTION

Clinical real-world data (RWD) are observational data describing patient health and health care delivery. RWD can be collected from a variety of sources, including electronic health records (EHRs), insurance claims databases, disease registries, and patient surveys, and can be a valuable source of real-world evidence (RWE) for studies on patient outcomes, investigational therapies, and clinical best practices.[1–4] Because RWE reflects clinical decision-making not constrained by a study protocol, it may better capture the true experience of patients living with a particular disease or receiving a particular therapeutic intervention, as compared to data collected through clinical trials or other restrictive clinical research settings. Further, RWE can be assembled from cohorts with greater diversity in patient backgrounds and geography, which can be especially useful for studies involving patients with rare disease. Given the potential utility of RWE in drug development, the U.S. Food and Drug Administration (FDA) has formally established guidance for using RWD in premarket and postmarket evaluations of novel treatments, including clinical studies to support regulatory approval decisions of new drugs or biological products, or new indications for existing ones.[5,6]

While both insurance claims and EHR data can provide patient health and care delivery data including diagnoses, prescriptions, and procedures, EHRs offer a richer and more nuanced source of RWD.[7] Radiographic images, genetic and molecular test results, pathology reports, and especially free text notes which may be part of the full medical record can contain highly valuable clinical insights not available in claims data. However, there are also several challenges with harnessing EHR data as a source of clinical evidence. Many components of EHRs, including the free-text notes from clinicians, are not standardized or structured, which can complicate automated extraction. Without automation, data extraction from EHRs requires time- and skill-intensive manual chart review by clinical experts and cannot be scaled to large cohorts. In addition, there are many EHR systems in use across healthcare entities in the U.S., which adds complexity to data management and utilization. Given that many patients switch health care systems or insurance providers frequently over time, an EHR-based dataset may have missing data if taken from a single healthcare system. Finally, almost all secondary data sources contain de-identified patient data, which disrupts longitudinal tracking across institutions and linking of patient-reported outcomes to disease course and treatment data.

To address some of these gaps, we set out to build a real-world data extraction platform that processes data from EHRs into structured and queryable data elements. Upon patient consent, medical records (digital or paper) from multiple institutions were collected and then analyzed by natural language processing algorithms to create structured data that was confirmed by clinical experts via two rounds of human review, confirming the data accuracy. Each patient participant had access to both their structured data and their complete collection of medical records, which they could use as they saw fit, e.g., second opinions, clinical trial matching. Collectively, the processed data could be used for retrospective, observational research including regulated studies on novel therapies. RWD from this platform (hereinafter “Invitae’s Ciitizen platform”) has already been used as natural history data to support a successful Investigational New Drug application for a novel treatment for pediatric patients with a severe form of epileptic encephalopathy [8].

The objective of the study presented here is to use the classic measures of precision and recall to validate the accuracy and comprehensiveness of Invitae’s Ciitizen platform performance as compared to manual chart review, the gold standard method for extracting data from medical records, in a cohort of metastatic breast cancer patients. A second objective is to demonstrate the potential for this data to generate clinical insights that may support hypotheses through a study of the time-to-distant metastasis (TDM) associated with specific tumor and patient characteristics.

## METHODS

### Overview of data extraction process

An overview of the data extraction process is shown in Figure 1. Following a directive from a consenting participant, the Health Insurance Portability and Accountability Act of 1996 (HIPAA) right of access is leveraged to obtain medical records from every institution reported to have been visited by the individual. A triage process guarantees recency of data and a minimum completeness of record collection. All medical records are stored electronically, and the participant can freely access them by logging into a secure portal. Medical record documents are then processed through natural language processing pipelines to convert unstructured data such as clinician notes and pathology reports into structured data consisting of clinical variables and their associated date(s) or date ranges that can be supported by multiple source documents (Figure 1). For example, a diagnosis of breast cancer on a particular date would be a single clinical variable that is supported by one or more medical records from multiple institutions. The automated extraction process includes document classification (e.g., pathology report, progress report, genetic testing results) and structuring of clinical terms to a pre-established dictionary of data variable types specific to solid tumors (Supplemental Table 1). In addition, modeling relationships connect certain variables, for example Adverse Events are modeled to the causative Medication, and Secondary Diagnoses (e.g., metastasis to bone) are modeled to the Primary Diagnosis (e.g., breast cancer). Following computational extraction, the structured data is reviewed by clinical experts who confirm clinical variables are accurate as supported by at least one source document for the term and associated date(s), as well as modeling relationships as relevant. The resulting data elements are stored securely in a HIPAA-compliant, controlled access, indexed database.

**Figure 1.**
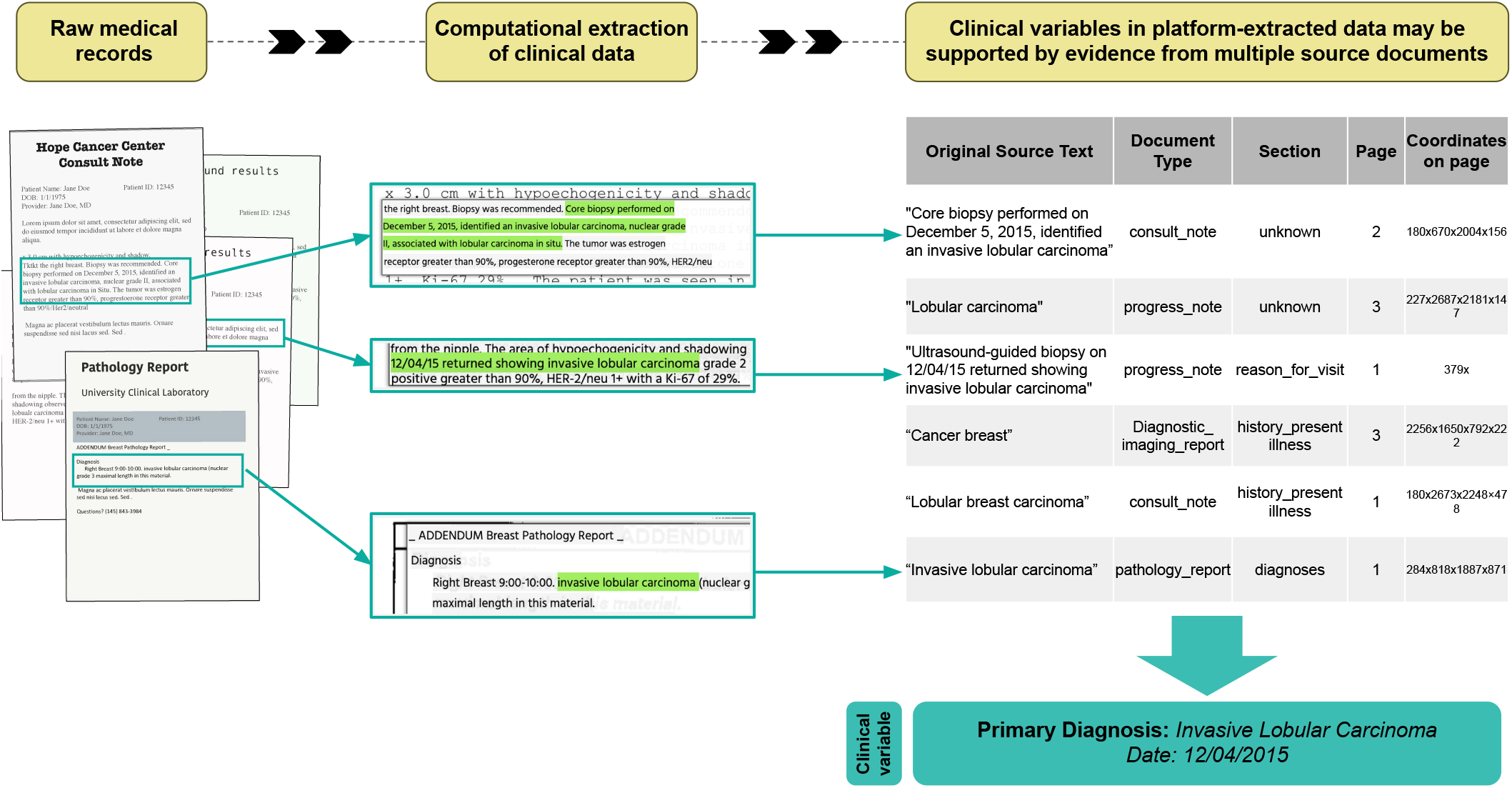
Platform Overview. Clinical data is computationally extracted from raw medical records and confirmed by human review based on one or more supporting documents.

### Validation study

#### Validation study design

This validation study protocol received a determination of exempt status by Pearl IRB. Fifty patients with metastatic breast cancer who had provided research consent to using their de-identified medical records data were randomly selected from our processed metastatic breast cancer cases to be the validation cohort. Data extracted from medical records of these 50 selected patients was compared to chart review conducted by oncology nurse annotators (Supplemental Figure 1), as in [9]. The oncology nurse annotators were contracted specifically for this analysis, in order to eliminate any bias due to familiarity with the extraction process or data structuring. Annotators completed electronic case report forms (CRFs) for 50 patients with metastatic breast cancer using either raw medical record documents (“records direct”) or the structured data produced by the platform. Variables on the CRF included primary breast cancer diagnosis, tumor histologic type, molecular type, stage, medications, disease statuses, and adverse events (Supplemental Table 1). Dates were required for most variables on the CRF, with the exception of comorbidities which were allowed to be undated. Each patient was reviewed by two different annotators, one who used the raw medical records and another who used the platform-extracted data.

#### Validation study analysis

Completed CRFs for the same patient were compared with the records-direct CRFs serving as the reference standard. The comparison was completed in two phases. In the first phase, variables identified in both the records-direct CRF and the platform CRF were designated as true positives (TPs), variables identified in the records-direct CRF only were considered false negatives (FNs), and variables identified in the platform CRF only were considered false positives (FP). However, as manual chart review is not necessarily error-proof, all FP and FN identified in the first phase were escalated to review by a third annotator. The escalation annotator reviewed source documents to determine if any variables in a platform CRF that were initially scored as FP could be verified in the source documents or extracted data and were therefore missed in manual chart review. If the variables were verified in the source documentation, the score was adjusted from FP to TP. For any variables scored FN because they were identified in the records-direct CRF but not the platform CRF, a third annotator determined if any were out of scope for the study, such as diagnostic procedures done before the primary diagnosis, which the platform does not extract. If so, the score was considered to be manual chart review error and adjusted from FN to null (not scored). These adjustments enabled a more accurate comparison between the records-direct and platform conditions.

Two metrics were calculated to assess the accuracy and comprehensiveness of the extracted data: precision, calculated as the number of TP divided by the sum of TP and FP; and recall, calculated as the number of TP divided by the sum of TP and FN.

### Demonstration of clinical utility

To assess the potential for the platform-extracted data to support clinical insights, we looked for correlations between clinical features and TDM among a cohort of patients with breast cancer from the platform database with distant metastasis (hereafter, the discovery cohort). From a starting population of 1,011 research-consented patients with breast cancer, we identified those with medical records data on the date of primary breast cancer diagnosis, date of documented metastasis to brain, bone, liver, or lung, and at least one of: germline *BRCA* status, HER2/HR molecular type, and histologic type. Separately, we examined the distribution of histologic types, molecular types, and stage at diagnosis among patients with invasive breast cancer in the Surveillance, Epidemiology, and End Results Program (SEER) cancer statistics registry (data from 1975–2017) to provide context for our platform-derived cohorts.

Demographics and clinical characteristics were assessed in the full study cohort. Clinical characteristics included histologic type (ductal, lobular, ductal + lobular, inflammatory, and metaplastic) and molecular type (based on combinations of hormone receptor positive [HR+], hormone receptor negative [HR-], human epidermal growth factor receptor 2 positive [HER2+], and human epidermal growth factor receptor 2 negative [HER2-]). The time to metastasis (TDM) was defined as the time between the first primary diagnosis and the first metastatic diagnosis at each site, e.g. metastasis to lung. For patients who had metastases to multiple sites, the first instance of metastasis to each site was calculated separately, in order to enable analyses based on site of disease. The average TDM was calculated across the discovery cohort and stratified by histologic type, molecular type, germline BRCA status, and site of distant metastasis. Bivariate differences in average TDM were determined by unpaired, two-tailed t-test with Welch’s correction. Multivariable analysis was performed using ANOVA and Tukey’s Honestly-Significant-Difference. Statistically significant findings were those with a p-value <0.05.

## RESULTS

### Validation Study: agreement between data extraction platform and manual chart review

Among the fifty patients with metastatic breast cancer in the validation cohort, the mean age at diagnosis was 47.7 (SD: 9.3; Range: 29–69), the most common histology was Invasive ductal carcinoma (62%), and the most common molecular type was HR+/HER2- (68%) (Table 1). Initial comparisons of CRFs completed with platform data and those completed directly from the medical records revealed 4,089 variables for comparison (Supplemental Table 2). Escalated review to validate or correct all false negatives (FNs) and false positive (FPs) by a third annotator revealed 798 variables that were either putative false positives (FPs) that were confirmed in the platform data source documents and missed in the manual chart review (and therefore re-coded as True Positives), or putative false negatives (FNs) that were outside of the scope of the study (e.g., a confirmed benign lump in the other breast prior to the primary diagnosis) and erroneously included in the records-direct CRF (Supplemental Table 2), which were then removed from scoring.

**Table 1.** Validation Cohort.

| **Characteristic** | **n=50** |
| --- | --- |
| **Age at diagnosis, Mean yrs (SD; range)** | 47.7 (9.3; 29–69) |
| **Histology, No. (%)** |  |
| Invasive ductal carcinoma | 31 (62%) |
| Invasive lobular carcinoma | 5 (10%) |
| Invasive ductal carcinoma and Invasive lobular carcinoma | 4 (8%) |
| Inflammatory | 2 (4%) |
| Metaplastic | 2 (4%) |
| Other / Unknown | 6 (12%) |
| **Molecular type, No. (%)** |  |
| HR+/HER2+ | 3 (6%) |
| HR+/HER2- | 34 (68%) |
| HR-/HER2+ | 4 (8%) |
| HR-/HER2- | 9 (18%) |
| Unknown | 0 (0%) |
| **Germline *BRCA* type, No. (%)** |  |
| *BRCA* Negative | 32 (64%) |
| *BRCA* Positive | 1 (2%) |
| Unknown | 17 (34%) |

After escalation and correction, 3,292 variables were available for comparison. As determined by the platform CRF, Invitae’s Ciitizen data extraction platform had an overall precision of 97.58% (2,619/2684) (Figure 2) when compared to manual chart review. When stratified by variable type, precision levels ranged from 91.98% (tumor status) to 100% (histologic type, grade, lab result, and most recent performance status).

**Figure 2.**
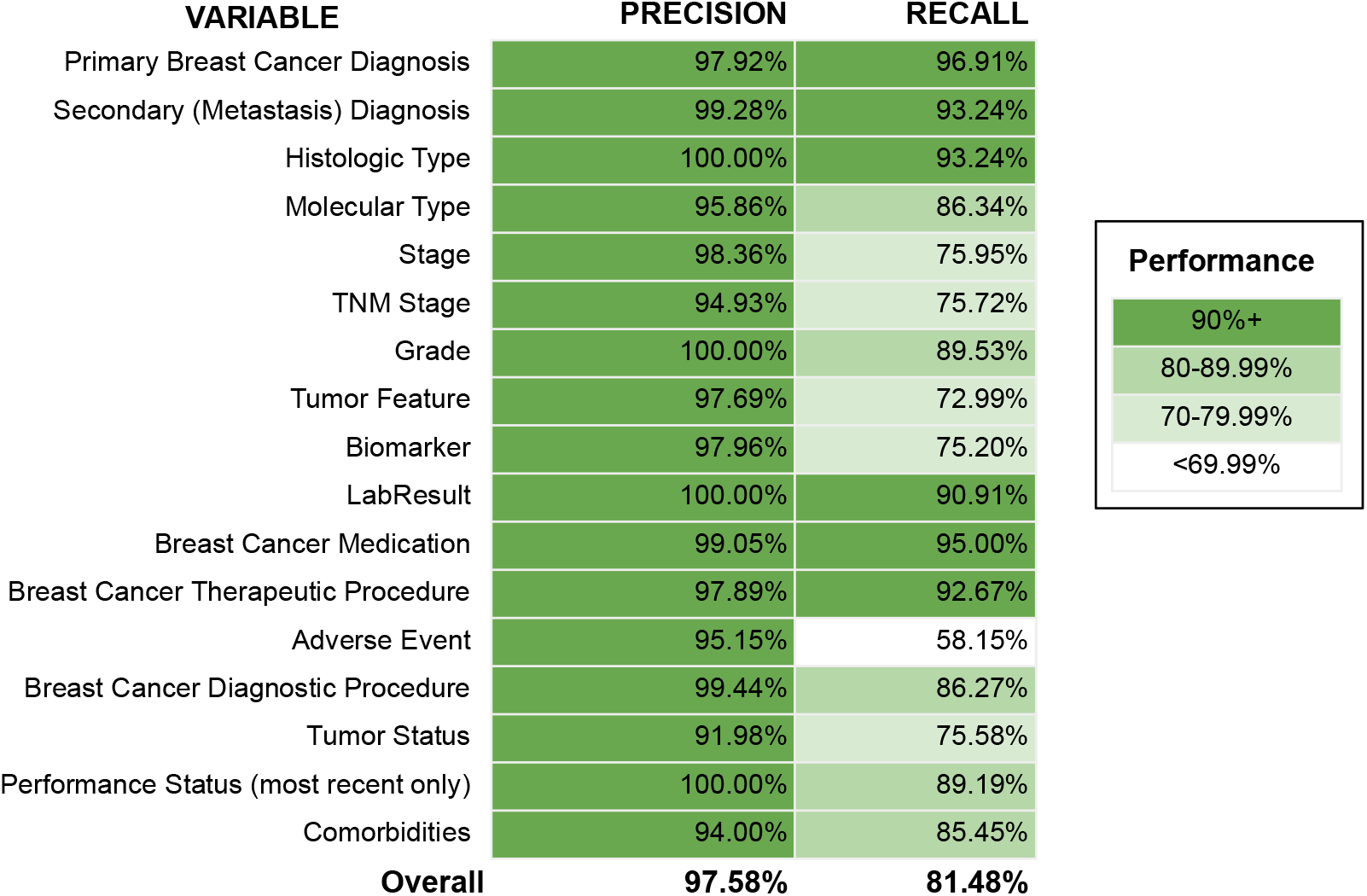
Performance of medical record extraction platform. Performance of the data-extraction platform was determined by comparing case report forms completed using platform-extracted data or medical record manual review. Variables identified in both the records-direct CRF and the platform CRF were designated as true positives (TPs), variables identified in the records-direct CRF only were considered false negatives (FNs), and variables identified in the platform CRF only were considered false positives (FP). Precision was calculated as the number of TP divided by the sum of TP and FP and recall was calculated as the number of TP divided by the sum of TP and FN.

The overall recall capability of the platform was 81.48% (2,618/3,213) (Figure 2). By individual variable, recall ranged from 58.15% (adverse events) to 95.00% (breast cancer medication) (Supplemental Table 2).

### Additional performance measures

The average time required to generate a CRF was 359.2 minutes (range 103–1,010 minutes; median 272.5 minutes) when medical records were manually reviewed and 49.4 minutes (range 14–130 minutes; median 45 minutes) when platform-extracted data was used. Thus, compared to manual chart review, completion of a CRF via extracted data was 7.3x faster when comparing average times and 6.1x faster when comparing median times.

The number of entities extracted by the platform varied by clinical variable (Supplemental Table 2) and reflects clinical expectations. Primary Diagnosis, which is captured as a single entity regardless of the duration, had a relatively low occurrence (mean of 2.2 per individual). Values over 1 are expected as this variable type includes not just the initial breast cancer diagnosis, but periods of NED and any recurrences. Conversely, entity types that are expected to occur multiple times during the patient journey had higher average numbers of entities per individual, for example anti-cancer medications (mean of 4.4) and tumor statuses (mean of 5.2). Biomarker data had the greatest number of entities, with an average of 12.7 instances of biomarker data available per patient, which reflects the presence of multiple tests and / or broad genomic panels with multiple results.

### Demonstration of clinical discovery

Separately, we explored the ability for platform-extracted data to reveal clinical insights. Within a cohort of 1,011 patients with breast cancer with a similar disease profile to those in the SEER database (Table 2), we identified 194 patients who developed distant metastases in the bone, brain, liver, or lung and had clinical data available prior to their metastatic breast cancer diagnosis. In this discovery cohort, the average age at diagnosis was 43 (SD: 9.3; Range: 29– 69) and most patients (73.9%, 144/194) had invasive ductal carcinoma (Table 2). Patients most frequently had the HR+/HER2- molecular type (62.1%; 121/194). A small percent (7.7%; 15 out of 193 individuals with documented BRCA results) of patients had a pathogenic germline variant in either *BRCA1* or *BRCA2* documented in their medical records. Metastases were most common in bone tissue (Supplemental Table 3). Metastatic tumors in bone and brain were more common in patients with HR+ molecular type compared to HR- molecular type (Supplemental Table 4).

**Table 2.** Characteristics of metastatic breast cancer patients in the discovery cohort and the SEER database.

| Characteristic | Breast cancer patients in SEER<sup>a</sup> database | Breast cancer patients in platform database (n=1,011) | Metastatic breast cancer patients in platform database (n=194) |
| --- | --- | --- | --- |
| Age at diagnosis, Mean (range) | NA | 47 (23–80) | 43 (26–73) |
| Histology, No. (%) |  |  |  |
| Invasive ductal | 319,963 (72.4) | 745 (74.3) | 144 (73.9) |
| carcinoma |  |  |  |
| Invasive lobular carcinoma | 43,930 (9.9) | 80 (8.0) | 17 (8.7) |
| Invasive ductal carcinoma and Invasive lobular carcinoma | 43,090 (9.7) | 32 (3.2) | 16 (8.2) |
| Metaplastic | NA | 36 (3.6) | 6 (4.6) |
| Papillary | 3,326 (0.8) | 4 (0.4) | 0 (0) |
| Other or unknown | NA | 114 (11.3) | 9 (4.6) |
| **Molecular type, No. (%)** |  |  |  |
| HR+/HER2+ | NA (10.0) | 128 (12.7) | 19 (9.7) |
| HR+/HER2- | NA (68.0) | 552 (54.9) | 121 (62.1) |
| HR-/HER2+ | NA (4.0) | 59 (6.0) | 10 (5.1) |
| HR-/HER2- | NA (10.0) | 152 (15.1) | 19 (9.74) |
| Unknown | NA (7.0) | 115 (11.3) | 26 (13.3) |

<sup>a</sup>Surveillance, Epidemiology, and End Results Program data from 1975–2017. For some SEER data, only percentages are available.
NA, not available

The average (range; SD) time to distant metastasis (TDM) was 2,295.1 (31–11,985; 2020.9) days across the discovery cohort. Patients with HR-/HER2- molecular type had the shortest average (SD) TDM (1187.4 [1,129.4] days), significantly shorter (p<0.0001) than those with the HR+/HER2- molecular type which had the longest (2,242.5 [2,042.5] days) (Figure 3A). When the histologic type of the tumor was considered, patients with ductal carcinomas had an average (SD) TDM of 2,270.1 (1,796.6) days, and those with lobular carcinomas had an average (SD) TDM of 2,812.0 (2,318.5) days (Figure 3B). Patients with the rarer metaplastic and inflammatory histologies had an average TDM of 739 (354.8) days and 1,005.8 days (166.2), respectively (Figure 3B), and these were both significantly shorter than the average TDMs for lobular and ductal types (p<0.005).

**FIGURE 3.**
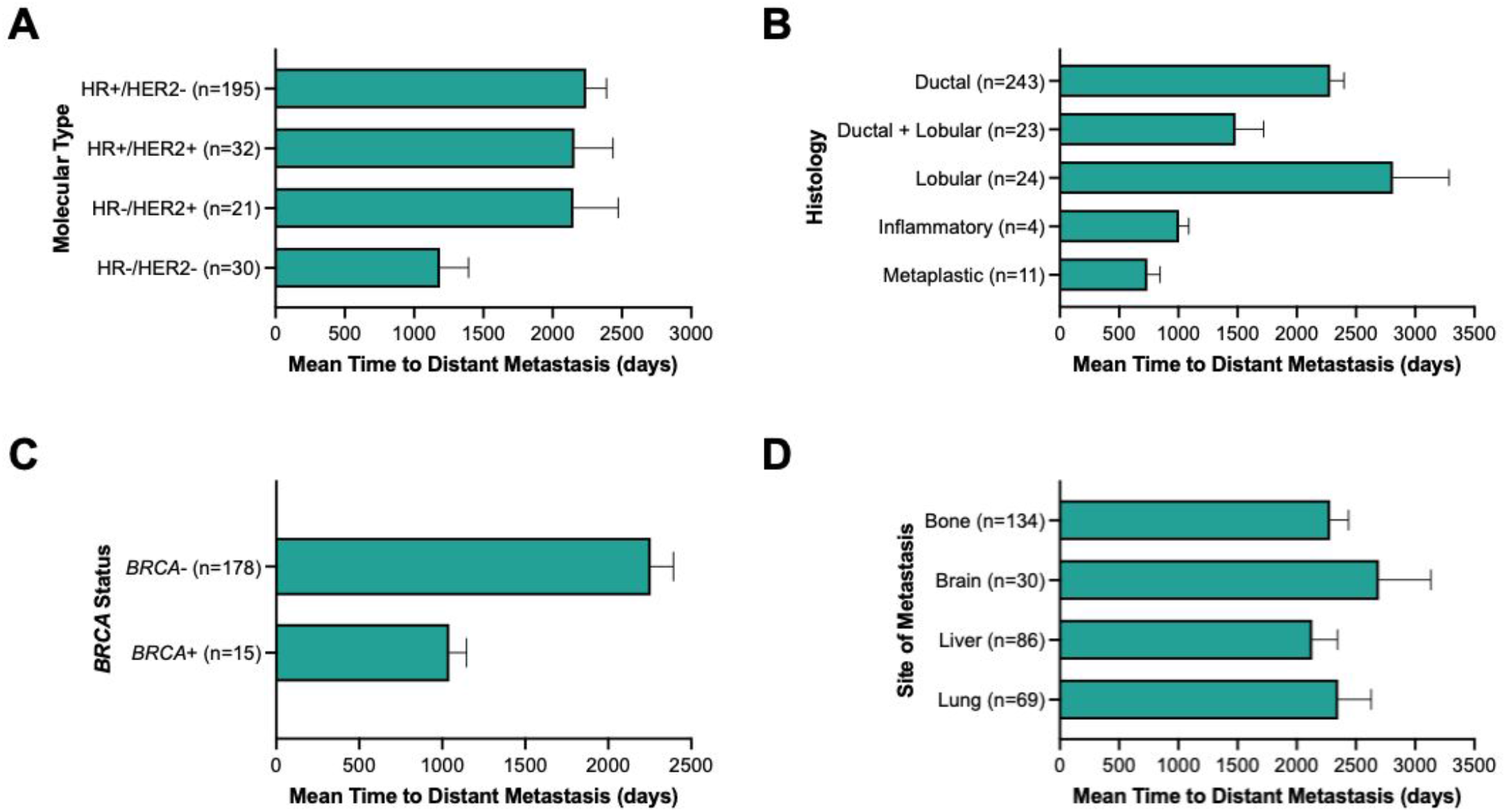
Tumor characteristics associated with shorter average time to distant metastasis. (A) molecular type, (B) histologic type, (C) *BRCA* status, and (D) location of metastasis. Error bars show standard error of the mean.

By *BRCA* germline status, individuals with pathogenic germline variants in genes *BRCA1* or *BRCA2* (or reported as “*BRCA* positive” in their medical records) had an average (SD) TDM of 1,043 (400.8) days, significantly shorter (*P*=0.0007) than individuals testing negative for *BRCA1/2* variants, who had an average (SD) TDM of 2,255.9 (1,822.6) days (Figure 3C). By site of distant metastasis, a liver metastasis was associated with the shortest average (SD) TDM at 2,131.4 (1,974.5) days and a brain metastasis was associated with the longest average (SD) TDM at 2,693.8 (2,407.6) days (Figure 3D).

In multivariate analyses of histology and molecular type, the shortest average TDM was observed among patients with combined ductal and lobular histologies and HR-/HER2+ molecular type (633 days), metaplastic histologic type and HR+/HER-2 molecular type (706 days) and metaplastic histologic type and HR-/HER- (Figure 4). By histology and site of metastasis, the shortest average TDMs were among patients with metaplastic histology and metastasis to bone tissue (522.5 days), combined ductal and lobular histology and metastasis to lung tissue (573.3 days), and metaplastic histology and metastasis to lung tissue (610.7) (Supplemental Table 4). By molecular type and site of metastasis, the shortest average TDMs were among patients with HR-/HER2- molecular type and metastasis to the lung (940.0 days), liver (999.5 days), and brain (1,217.0 days). Finally, patients who were *BRCA* positive had shorter average TDM for all sites of metastasis (Supplemental Table 4).

**FIGURE 4.**
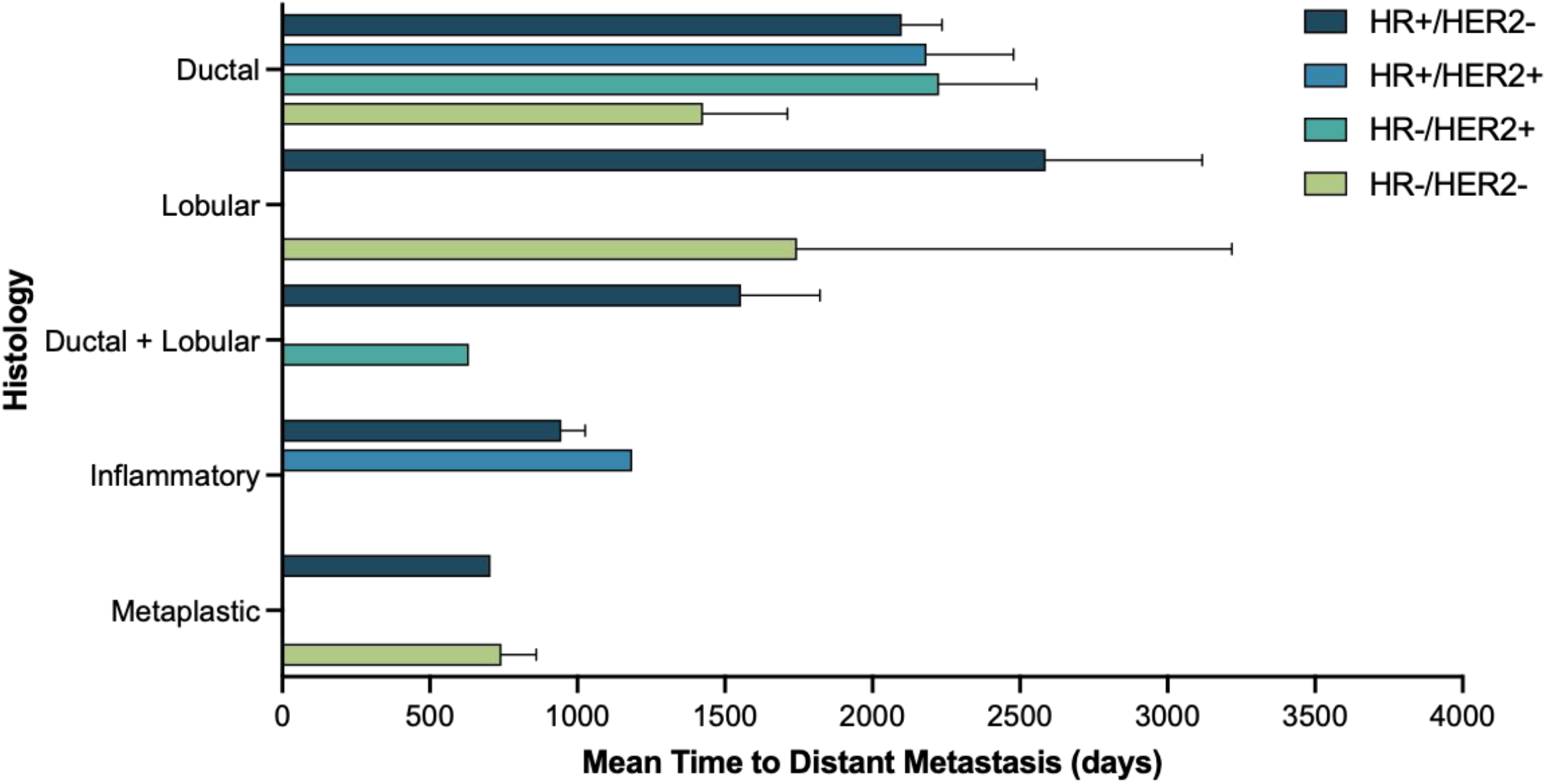
Multivariable analyses of tumor characteristics with time to distant metastasis. Error bars show standard error of the mean.

## DISCUSSION

In this validation and demonstration study, we showed that a novel EHR data-extraction platform can reliably identify clinical variables in medical records, including from unstructured data in clinician notes and pathology reports, with a high level of precision (97.58%) and recall (81.48%). The platform-extracted data is also relevant to the study population of patients with metastatic breast cancer as shown through our comparison to the SEER registry. Finally, we demonstrated how this platform-extracted RWD can provide clinical insights, as the platform-extracted data revealed statistically significant associations between tumor characteristics and the average TDM for patients with metastatic breast cancer.

In addition to its utility for regulatory applications, the clinical data collected by the platform can also be used by patients to better track their diagnosis and treatment history. Patients living with cancer or rare disease face serious information burdens, multiple clinic or hospital visits, challenging terminology, and complex treatment options,[10]. A prior study has shown that nearly all patients appreciate greater access to their clinical information,[11] thus the patient-centered approach of the platform described here could be a solution to this issue.

RWD has long been used to assess drug safety, but now has the potential to address significant gaps in drug development.[12] New medicines and other medical products regulated by the US government are largely tested through clinical trials in controlled settings that are expensive and cumbersome and may not fully reflect the real-world experience of individuals who may one day use those products. In addition, clinical trials are typically restricted to regions with academic medical centers where the patient population may not reflect the true diversity of individuals affected by a disease.

RWD can come from many sources, including insurance claims data, epidemiological survey data, patient reported outcomes, and aggregated EHR data. Claims data can provide longitudinal information about individuals who are continuously enrolled in specific health insurance plans, but has been shown to lack details on clinical variables, interventions, and outcomes sufficient for research use,[13] and will likely have missing data if individuals change insurance providers or programs.[7] Epidemiological survey data such as the SEER database have limited treatment information and capture only a snapshot of a given individual’s health, thereby losing longitudinal information. EHRs contain detailed and longitudinal information about a patient’s care, and may even include diagnosis and procedure codes found in claims data but can have major shortcomings. For example, the clinical notes written by healthcare providers may contain a wealth of information but the unstructured format of this data type renders it nearly useless for large cohort analyses. However, others have shown that methods for computationally analyzing free text data can identify patient outcomes such as cancer metastases and other cancer events as well as adverse drug events.[2,12,13] Another potential shortcoming of EHRs is the fact that they are typically siloed to individual institutions precluding longitudinal study of patients who receive care at multiple institutions, resulting in significant gaps in a patient’s treatment journey over time.

The Invitae Ciitizen EHR data-extraction platform described in this study can resolve these shortcomings, as it draws records from all institutions identified by a patient and uses computational methods to identify and give structure to clinical variables, which are all confirmed by oncology nurse annotators. An additional advantage of the platform is the close involvement with patients, whose involvement begins with their personal directive for platform experts to request records from any institution where they have received care, and may include permission to be re-contacted for follow-up surveys (with financial compensation) that may address chronic symptoms and quality of life. Patients also have the ability to share their data with other clinicians for second opinions and with clinical trial matching services. Although this study focused on patients with breast cancer, the clinical-extraction platform supports medical record extraction for several other solid cancer types, hematologic malignancies, and neurodevelopmental disorders and has the capability to expand and accommodate any health condition.

While the precision of all variables in this study was uniformly high (94.00%–100.00%), we observed a wide range of recall performance across variables in the validation study (58.15%– 96.91%). In particular, adverse events had only 58.15% recall, which may reflect challenges for the natural language processing algorithm to identify causative statements.

In addition to its potential utility as a source of RWE for regulatory filings, the Invitae Ciitizen medical record-extraction platform can reveal clinical insights as demonstrated by our analyses of TDM in breast cancer patients. While it has been previously demonstrated that HR+/HER2- breast cancer is less likely to spread to the brain than the other subtypes,[14] and that metaplastic breast cancer is more likely to be HR-/HER2-,[15] our analyses also revealed some novel findings that could inform more personalized screening recommendations for patients with breast cancer. In particular, individuals with the shortest average TDMs included those with metaplastic and inflammatory subtypes, HR-/HER- tumor molecular types, and germline variants in *BRCA1* or *BRCA2*. Our findings suggest that histology, molecular type, and genetic risk factors may be worth considering when selecting the frequency and timing of screening.

There are limitations to this study worthy of consideration. Given that our data set is based on real-world clinical experience of individuals and accommodates more than one variable per type (for example, any one individual may have zero to numerous medications), it was not possible to establish true negatives in our performance analyses and therefore we could not assess specificity. In addition, our validation was conducted with a small sample size (n=50) due to the intensive personnel requirements for conducting manual chart reviews. Finally, our validation study does not account for source text quality as clinical variables may be sourced from any type of document (see Figure 1), and medical records (and the structured data extracted from them) may not contain all the relevant medical and patient information.

## CONCLUSION

An EHR data-extraction platform can produce structured datasets with high precision and recall and the resulting data can be used as RWD in regulatory filings or clinical discovery. Given that the platform uses disease-specific models, future efforts should validate the platform in other oncology and non-oncology patient groups.

## Data Availability

The data underlying this article cannot be shared publicly due to the privacy of individuals that participated in the study. De-identified data will be shared on reasonable request to the corresponding author.

## CONTRIBUTORS

This study was performed under the supervision of authors AB, AN, and LWO, who provided significant contributions toward its design and implementation. Authors AN, SA, LH, HS, and JT conducted the manual medical records reviews and completed the case report forms. Authors AN, AC, HEL, LWO did the discovery cohort analyses. AN and SR drafted the manuscript, AN, SR, and EB generated figures and tables, and all authors provided critical review of the manuscript.

## ACKNOWLEDGMENTS

The authors thank Robert L. Nussbaum, Deven McGraw, Flavia Facio, and Luc Cary for their thoughtful review and comments on this manuscript. In addition, we thank the patients who contributed their data for research.

