## Supplemental Data for "Validation and clinical discovery demonstration of a real-world data extraction platform"

**Supplemental Figure 1. Overview of validation study methods**

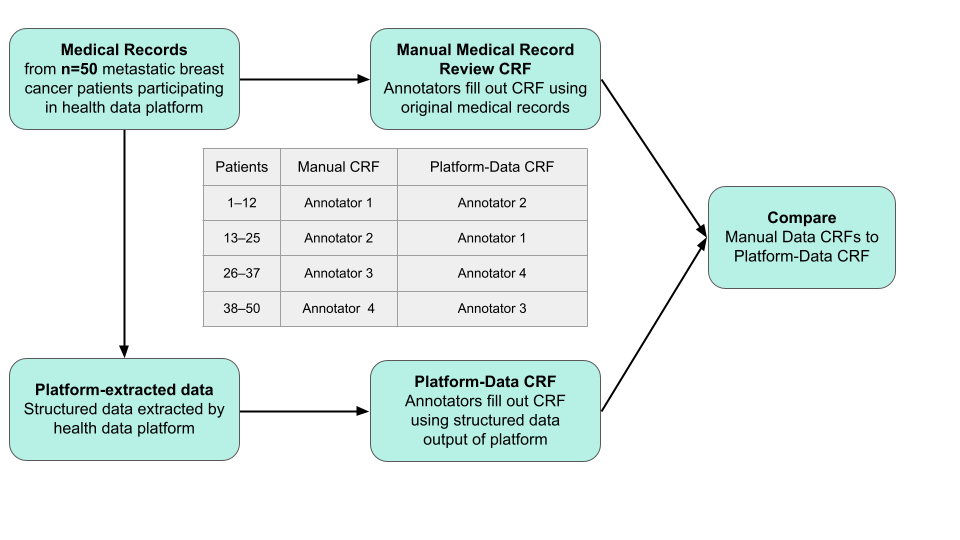

CRF, case report form.

**Supplemental Table 1. Variables on the case report form.**

| **VARIABLE** | **FORM VARIABLE CONSTRAINTS** | **OTHER ATTRIBUTES** |
| --- | --- | --- |
| UUID | Pre-populated |  |
| BiologicalSex | Female, Male |  |
| ZipCode | 5 digit number |  |
| MonthofBirth | 2 digit month, 4 digit year |  |
| Primary Breast Cancer Diagnosis | Dropdown list | Event Date, Earliest Date, Latest Date |
| Secondary (Metastasis) Diagnosis | Dropdown list | Event Date  Modeling to Diagnosis |
| HistologicType | Dropdown list | Event Date  Modeling to (Secondary) Diagnosis |
| MolecularType | Dropdown list | Event Date  Modeling to (Secondary) Diagnosis |
| Stage | Dropdown list | Event Date  Modeling to Diagnosis |
| TNMStage | Dropdown list | Event Date  Modeling to Diagnosis |
| Grade | Dropdown list | Event Date  Modeling to (Secondary) Diagnosis |
| TumorFeature | Dropdown list | Event Date  Modeling to (Secondary) Diagnosis |
| Biomarker | Dropdown list | Event Date  Panel / Test Name  Method  Variant  Result / Value  Modeling to (Secondary) Diagnosis |
| LabResults | Dropdown list | Event Date  Result / Value  Unit  Range |
| Breast Cancer Medication | Dropdown list | Event Date, Earliest Date, Latest Date |
| Breast Cancer TherapeuticProcedure | Dropdown list | Event Date, Earliest Date, Latest Date  Modeling to (Secondary) Diagnosis |
| Breast Cancer Diagnostic Procedure | Dropdown list | Event Date  Modeling to (Secondary) Diagnosis |
| Tumor Status | Dropdown list | Event Date, Earliest Date, Latest Date  Modeling to (Secondary) Diagnosis |
| Comorbidity | Dropdown list | Event Date, Earliest Date, Latest Date |
| Adverse Event | Dropdown list | Event Date, Earliest Date, Latest Date  Modeling to Medication or TherapeuticProcedure |
| PerformanceStatus (most recent only) | Dropdown list | Event Date |

**Supplemental Table 2.** Variables in validation study, outcome of escalated review, and platform performance by variable

|  | **No. of truth set variables (Mean number per patient, N=50)** | **Phase 1 review** | | **Final validation variables** | | | **Performance of platform CRF by variable** | |
| --- | --- | --- | --- | --- | --- | --- | --- | --- |
| **Variable** |  | **No. variables reviewed** | **No. variables removed in escalated review** | **True positive** | **False negative** | **False positive** | **Precision TP/(TP+FP)** | **Recall**  **TP/(TP+FN)** |
| Adverse Event | 270 (5.4) | 306 | 28 | 157 | 113 | 8 | 95.15 | 58.15 |
| Biomarker | 637 (12.74) | 796 | 149 | 479 | 158 | 10 | 97.96 | 75.20 |
| Breast Cancer Diagnostic Procedure | 415 (8.3) | 808 | 391 | 358 | 57 | 2 | 99.44 | 86.27 |
| Breast Cancer Medication | 220 (4.4) | 235 | 13 | 209 | 11 | 2 | 99.05 | 95.00 |
| Breast Cancer Therapeutic Procedure | 150 (3.0) | 197 | 44 | 139 | 11 | 3 | 97.89 | 92.67 |
| Comorbidities | 55 (1.1) | 70 | 12 | 47 | 8 | 3 | 94.00 | 85.45 |
| Grade | 86 (1.7) | 99 | 13 | 77 | 9 | 0 | 100.00 | 89.53 |
| Histologic Type | 222 (4.44) | 238 | 16 | 207 | 15 | 0 | 100.00 | 93.24 |
| Lab Result | 33 (0.7) | 47 | 14 | 30 | 3 | 0 | 100.00 | 90.91 |
| Molecular Type | 161 (3.2) | 188 | 21 | 139 | 22 | 6 | 95.86 | 86.34 |
| Performance Status (most recent only) | 35 (0.7) | 46 | 11 | 31 | 4 | 0 | 100.00 | 88.57 |
| Primary Breast Cancer Diagnosis | 110 (2.2) | 120 | 8 | 94 | 3 | 2 | 97.92 | 96.91 |
| Secondary (Metastasis) Diagnosis | 148 (3.0) | 161 | 12 | 138 | 10 | 1 | 99.28 | 93.24 |
| Stage | 79 (1.6) | 88 | 8 | 60 | 19 | 1 | 98.36 | 75.95 |
| TNM Stage | 173 (3.5) | 199 | 19 | 131 | 42 | 7 | 94.93 | 75.72 |
| Tumor Feature | 174 (3.5) | 193 | 16 | 127 | 47 | 3 | 97.69 | 72.99 |
| Tumor Status | 258 (5.2) | 298 | 23 | 195 | 63 | 17 | 91.98 | 75.58 |
| Total | 3226 | 4089 | 798 | 2618 | 595 | 65 | 97.58 | 81.48 |

**Supplemental Table 3.** Number of patients in discovery cohort by site of metastasis, histologic type, and molecular type.

|  | **Bone**  **(No. patients)** | **Brain**  **(No. patients)** | **Liver**  **(No. patients)** | **Lung**  **(No. patients)** | **All sites of metastasis**  **(No. patients)** |
| --- | --- | --- | --- | --- | --- |
| **Histologic type** | | | | | |
| Ductal | 98 | 25 | 63 | 56 | 242 |
| Ductal + Lobular | 13 | 0 | 7 | 1 | 23 |
| Lobular | 14 | 0 | 9 | 1 | 24 |
| Inflammatory | 1 | 0 | 1 | 2 | 3 |
| Metaplastic | 4 | 2 | 2 | 3 | 10 |
| All | 130 | 27 | 82 | 10 | 302 |
| **Molecular type** | | | | | |
| HR+/HER2- | 88 | 10 | 53 | 44 | 195 |
| HR+/HER2+ | 13 | 6 | 7 | 6 | 32 |
| HR-/HER+ | 5 | 5 | 7 | 4 | 21 |
| HR-/HER2- | 11 | 4 | 7 | 8 | 30 |
| All | 117 | 25 | 74 | 62 | 280 |
| **BRCA status** | | | | | |
| BRCA positive | 5 | 1 | 5 | 4 | 15 |
| BRCA negative | 76 | 17 | 44 | 41 | 178 |
| All | 81 | 18 | 49 | 45 | 193 |

HR, hormone receptor; HER2, human epidermal growth factor receptor 2

For patients with metastases to multiple sites, the earliest indication of a metastasis to each site (bone, brain, liver, or lung) is counted independently. BRCA status was based on lab test results in medical record or free text comment on patient’s BRCA status; patients without information on BRCA status were excluded.

**Supplemental Table 4.** Average time to distant metastasis among patients in the discovery cohort by site of metastasis, histologic type, and molecular type (N=194).

|  | **Bone** | **Brain** | **Liver** | **Lung** |
| --- | --- | --- | --- | --- |
| **Histologic type** | | | | |
| Ductal | 2376.3 | 2474.6 | 1955.3 | 2346.9 |
| Ductal + Lobular | 1461.7 | NA | 1914.7 | 573.3 |
| Lobular | 2874.1 | NA | 2830.6 | 1774.0 |
| Inflammatory | 868.0 | NA | 1186.0 | 984.5 |
| Metaplastic | 522.5 | 1001.0 | 1102.5 | 610.7 |
| **Molecular type** | | | | |
| HR+/HER2- | 2157.8 | 3084.9 | 2136.3 | 2348.6 |
| HR+/HER2+ | 2093.1 | 3044.7 | 2105.3 | 1476.5 |
| HR-/HER+ | 2423.4 | 2058.2 | 2144.0 | 1936.3 |
| HR-/HER2- | 1267.6 | 1217.0 | 999.5 | 940.0 |
| **BRCA status** | | | | |
| BRCA positive | 871.8 | 1035.0 | 1116.0 | 1166.0 |
| BRCA negative | 2360.0 | 2312.6 | 1970.4 | 2344.0 |

NA, not applicable (no patients in category); HR, hormone receptor; HER2; human epidermal growth factor receptor 2

For patients with metastases to multiple sites, the earliest indication of a metastasis to each site (bone, brain, liver, or lung) is counted independently. BRCA status is based on lab test result in medical record or free text comment on patient’s BRCA status; patients without information on BRCA status are included in the BRCA negative category.
