## Supplemental Figure 1 for "Validation and clinical discovery demonstration of a real-world data extraction platform"

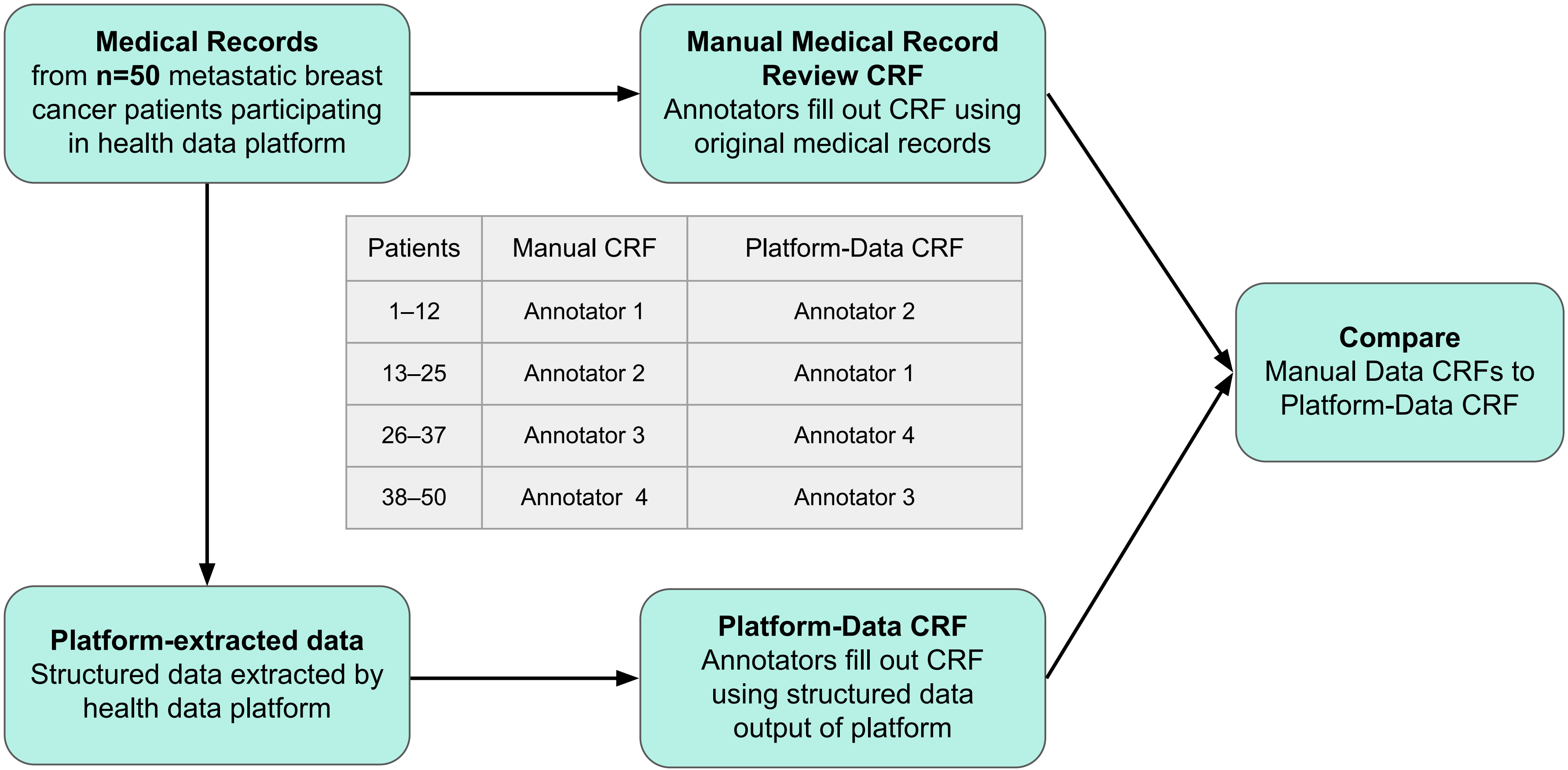


| Patients | Manual CRF | Platform-Data CRF |
|---|---|---|
| 1–12 | Annotator 1 | Annotator 2 |
| 13–25 | Annotator 2 | Annotator 1 |
| 26–37 | Annotator 3 | Annotator 4 |
| 38–50 | Annotator 4 | Annotator 3 |
